# MedKLIP: Medical Knowledge Enhanced Language-Image Pre-Training

**DOI:** 10.1101/2023.01.10.23284412

**Authors:** Chaoyi Wu, Xiaoman Zhang, Ya Zhang, Yanfeng Wang, Weidi Xie

## Abstract

In this paper, we consider the problem of enhancing self-supervised visual-language pre-training (VLP) with medical-specific knowledge, by exploiting the paired image-text reports from the radiological daily practice. In particular, we make the following contributions: **First**, unlike existing works that directly process the raw reports, we adopt a novel report filter to extract the medical entities, avoiding unnecessary complexity from language grammar and enhancing the supervision signals; **Second**, we propose a novel entity embedding module by querying an external knowledge description base, to exploit the rich context of additional information that the medical domain affords, and implicitly build relationships between entities in the language embedding space; **Third**, we propose a novel Transformer-based fusion model for spatially aligning the entity description with visual signals at the image patch level only with self-supervised learning, thus enabling the ability for spatial grounding; **Fourth**, we conduct thorough experiments to validate the effectiveness of our proposed architecture, and benchmark on numerous public benchmarks e.g., ChestX-ray14, RSNA Pneumonia, SIIM-ACR Pneumothorax, COVIDx CXR-2, COVID Rural, and EdemaSeverity. In both zero-shot and fine-tuning settings, our model has demonstrated strong performance compared with the former methods on disease classification and grounding.

## 1. Introduction

With the rapid development of deep learning, numerous works have been proposed to facilitate computer-aided diagnosis in the medical field [16, 17, 39, 48]. Despite the tremendous progress, these models are normally trained to recognize or segment the structures that fall into a certain closed set of anatomical or disease categories, whenever a new disease comes to be of interest, a costly procedure for data annotation, model re-training, and ethics proof will be required, fundamentally limiting its practical values. As an alternative, recent research considers to train the model by exploiting a large number of multi-modal medical data, that is generated from daily clinical routine, for example, the most common example is the dataset of X-ray images with paired radiological reports [15, 25, 28].

This paper focuses on self-supervised vision-language representation learning in the medical domain, with the goal of zero-shot disease diagnosis (classification) and grounding. Undoubtedly, such tasks have also been widely investigated in the computer vision community, with significant progress made in the past years, for example, CLIP [43], ALBEF [30], BLIP [29], etc. However, to achieve such a goal in the medical domain, different challenges must be resolved, which requires research efforts from the community: *First*, data availability, training Foundation Models in computer vision normally require over millions of image-text pairs at ease, while in the medical domain, only a few hundred thousand pairs are available [28]. The limited data amount challenges language models to understand the reports in free form [7]. *Second*, the problem considered in medical diagnosis is naturally fine-grained, that requires distinguishing between fine appearance details to understand the disease, as a consequence, domain knowledge is essential; *Third*, robustness is crucial, it is, therefore, preferable to have explainability, where diagnosis results come along with the visual grounding, to help radiologists to understand the system, and build trust between humans and machines.

Unlike existing work in medical VLP (Vision-Language Pre-training) [7, 22, 40, 56] that naïvely matches raw reports with image scans, we propose a novel knowledge-enhanced visual-language model that takes medical prior into consideration and enables us to address the aforementioned challenges explicitly, as shown in Fig. 1: *First*, we propose a report filter to extract useful medical entities, and simplify each report into sets of triplets, denoted as {entity, position, exist}. Consequently, decomposing reports into entities leads to an effective representation of the reports with minimal information loss, enriching supervision signals at the detailed entity level; *Second*, we map these entities into fine-grained descriptions by querying a well-defined medical knowledge base, and compute the text embedding for these descriptions, to implicitly build relationships between entities; *Third*, we adopt a transformer-based architecture for aligning the image patches with entity descriptions, that simultaneously infer the likelihood of certain diseases and the visual evidence in the form of a spatial heatmap, *i.e*., providing grounding for explainability purpose.

**Figure 1.**
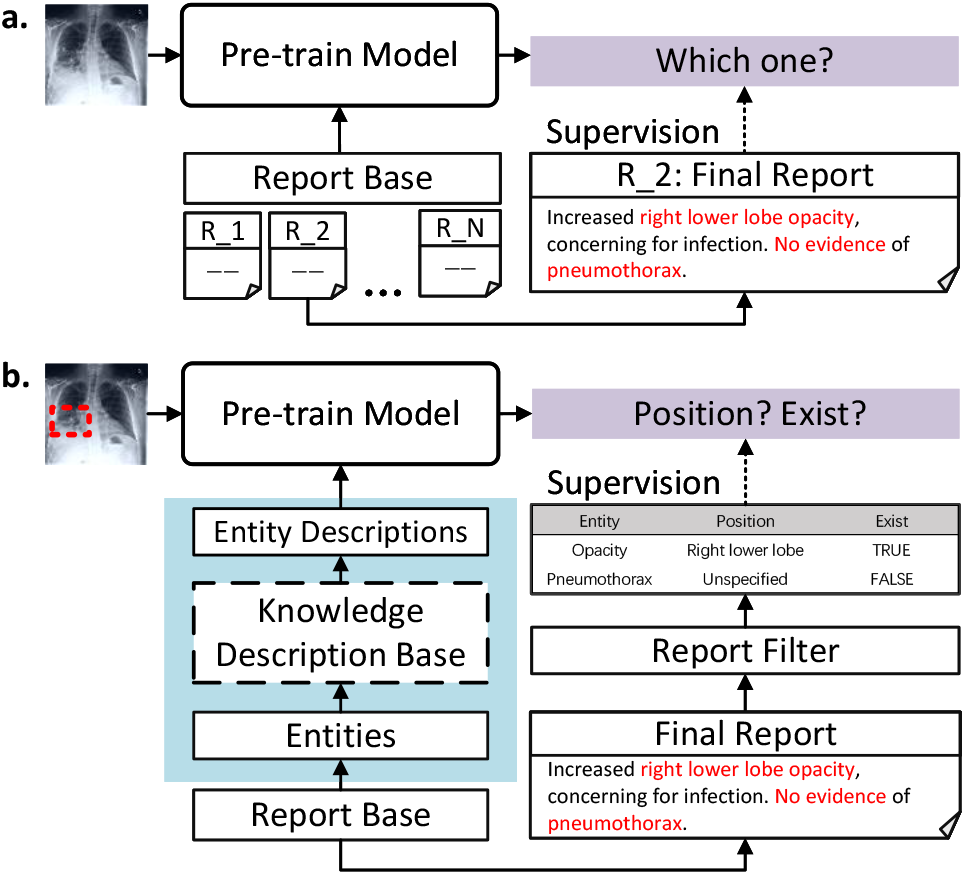
Our method mainly considers combining medical knowledge with VLP. **a**. shows the standard VLP flowchart which uses text-image retrieval as a proxy task. **b**. is our MedKLIP flowchart. We adopt a report filter module to decompose raw reports at entity level and further use knowledge descriptions to explain entities. Our model can realize zero-shot classification and grounding.

We train the model on the most widely-used medical image-report dataset MIMIC-CXR [28] and rigorously evaluate on numerous public benchmarks, *e.g*., ChestXray14 [50], RSNA Pneumonia [44], SIIM-ACR Pneumothorax [1], COVIDx CXR-2 [41], COVID Rural [12, 47], and EdemaSeverity [8]. We get a state-of-the-art zero-shot classification and grounding performance on different diseases, spanning different image distributions, with further fine-tuning, our model still exceeds previous models significantly.

## 2. Related Work

### Vision-Language Pre-training Models

Vision-Language Pre-training (VLP) models have achieved great success in natural scenarios. Generally, there are two typical structures for VLP models. One is two-stream methods [5,27,30]. The other is single-stream methods [10, 32]. These impressive results promote VLP methods in medical. Different from natural data, medical VLP suffers from a serious Lack of Data (224k [28] vs 400M [5]). Most medical VLP methods follow the two-stream methods [8, 22, 40, 56]. ConVIRT [56] first proposed to use contrastive learning as a proxy task in medical. LoVT and GLoRIA then focus on improving the local alignment performance [8, 22]. BioViL notices the language pattern in reports is different from other natural texts and re-designs the language model used for medical VLP [7]. These works have greatly contributed to the development of this aspect. However, they still view medical texts and images as common natural data and use a classical pipeline to handle them, instead of leveraging the rich prior knowledge in medical.

### Medical Named-Entity-Recognition Models

Various natural language processing (NLP) approaches have been proposed to extract useful information from radiology reports [25, 37, 42, 45]. These early methods considered only the disease, causing they lose a lot of information. Some Analysis tools [4,6] have also been developed to extract key clinical concepts and their attributes from biomedical text. Further state-of-the-art works [26, 51] are proposed to extract relationship between different entities without demand of pre-defined close disease set, retaining most of useful information with high accuracy. This progress inspires us a lot and provide a new perspective for VLP. However, how to take advantage of this Named-Entity-Recognition (NER) models have not been discussed sufficiently in VLP field.

### Medical Knowledge Enhanced Models

Leveraging external medical knowledge to enhance deep learning models is not a new topic [52]. Depending the approaches of using medical knowledge, They can be classified into modelbased and input-based two types. In model-based types, the authors may imitate the radiological practice to design the model [20, 23, 31, 49] or change the model structure based on diagnostic patterns [11, 18, 38]. In input-based types, the knowledge is viewed as an extra input to calculate features [46, 53, 54] or to guide the final loss [9, 24]. Multi-task learning and attention mechanism [33, 34, 36] are often adopted to realize medical knowledge combination. However, most of these works are task-specific [52] because medical knowledge lacks an effective shared representation space across various diseases while we demonstrate that leveraging medical entity descriptions with text encoding has the potential to provide such a space.

## 3. Method

In this section, we start by describing our considered problem scenario in Sec. 3.1, followed by the details of our proposed Transformer-based architecture in Sec. 3.2, including, the visual encoder, knowledge-enhanced text encoder, and the fusion module for aligning the visual and language signals. In Sec. 3.3, we describe the training procedure for our proposed model with the paired image-reports sourced from the daily routine X-ray scans.

### 3.1. Problem Scenario

Assuming we are given a training set with *N* samples, *i.e*., 𝒟_train_ = (𝒳_1_, *𝒯*_1_), …, (𝒳_*N*_, *𝒯*_*N*_), where 𝒳_*i*_, *𝒯*_*i*_ refer to the X-ray image and its corresponding medical report generated in the daily routine scans, respectively, our goal is to train a visual-language model that enables us to diagnose the existence of certain diseases and localize the visual evidence spatially. Specifically, at inference time, we can freely ask the system to identify the likelihood of the patient getting a certain disease (may or may not be seen during training), with its visual description for the disease of interest:

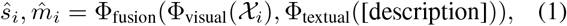

where 𝒳_*i*_ ∈ ℝ^*H*×*W* ×3^ refers to an image sample from the **test set**, with *H, W* denoting height and width respectively. ŝ_*i*_ ∈ [0, 1] refers to the inferred likelihood of the patient having a certain disease indicated by the input description, and 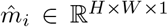 denotes a predicted spatial heatmap, with high activation on pixels that potentially provide the visual indication for such disease. In the following section, we will detail the individual components of our architecture, namely, the visual encoder, text encoder, and fusion module, and training them with the available training set (𝒟_train_).

### 3.2. Architecture

In this section, we detail our proposed framework, consisting of three main components, namely, visual encoding, knowledge-enhanced language encoding, and fusion module, as shown in Fig. 2. Note that, we hereon only consider single sampled image-reports pair (𝒳_*i*_, *𝒯*_*i*_), and ignore the subscript in notations for simplicity.

**Figure 2.**
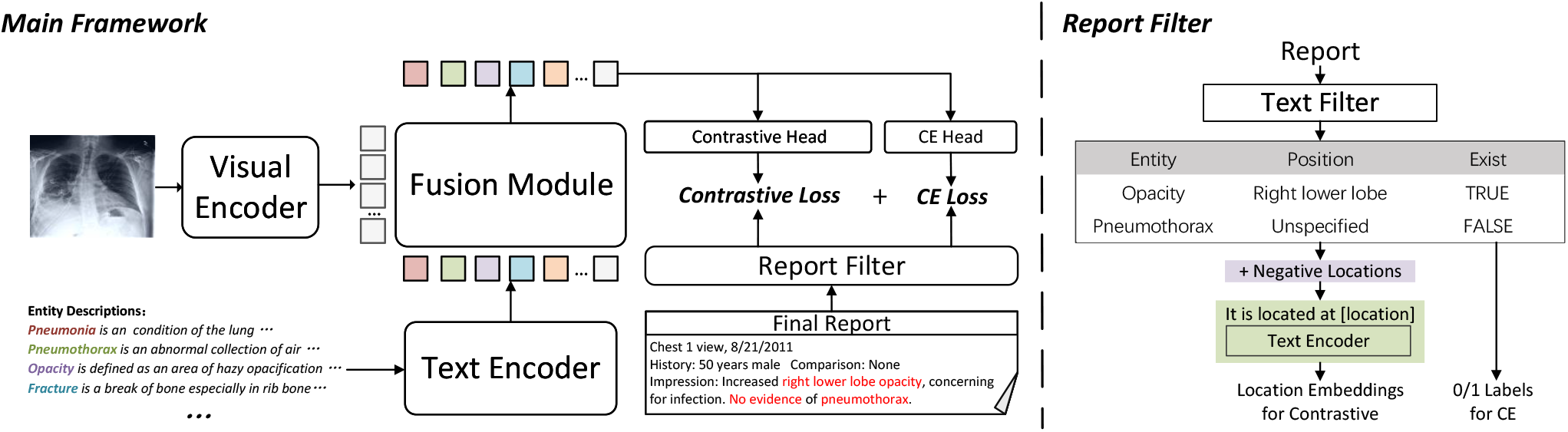
The whole framework of our method. The figure mainly contains the fourth module: *Visual Encoder, Knowledge-enhanced Language Encoding, Fusion Module. Knowledge-enhanced Language Encoding* contains *Text Encoder* and *Report Filter. Report Filter* extracts entities from the raw reports and *Text Encoder* further embeds them. *Visual Encoder* is used to encoder the input of visual modalities and *Fusion Module* is used for cross-modality interaction. The details of *Report Filter* can be found in the right sub-figure. A report is first filtered by a pre-trained filter and viewed as a set of triplets. The *“Position”* part is mixed with some negative positions for contrastive loss and the *“Exist”* part is used for CE loss.

#### 3.2.1. Visual Encoding

Given an X-ray image scan 𝒳 ∈ ℝ^*H*×*W* ×3^, we can compute the features with a visual backbone:

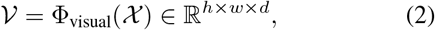

*h, w, d* refer to the height, width, and feature dimension of the output feature map, in our case, we adopt a standard ResNet-50 as the visual backbone, and take the output from the 4th residual block. Note that, we make the such an architectural choice for a fair comparison with existing work [7,22,40,56], while other visual backbones, *e.g*., ViT [14], can equally be applied.

#### 3.2.2. Knowledge-enhanced Language Encoding

The goal of this module is to extract useful information from the text report, by incorporating medical domain knowledge. In particular, we design two stages, namely report filtering, and entity encoding.

##### Report Filtering

To start with, we propose to condense the report and transform it into a set of entity triplets, *i.e*., removing the unnecessary complexity from language grammar, as shown in Figure 2 (right). In detail, we use a pretrained text filter [26, 55] to extract valuable information from the report, for example, the medical entities and their corresponding positions on the image.

Specifically, given a report 𝒯 with a set of sentences, 𝒯= {*s*_1_, *s*_2_, …, *s*_*M*_}, the filter independently operates on each of the sentences, and construct a number of triplets from the report, with the extracted entity (most are diseases), spatial position, and a label indicating the existence of the disease:

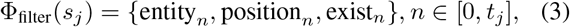

where *t*_*j*_ represents the total number of entities contained in one sentence, with *n* = 0 indicating the special case that there is no valid entity. **Note that**, the position refers to the spatial position of the entity lying in an image, it is not to be confused with the positional embedding in Transformer.

##### Entity Encoding

Here, we replace the entities by querying detailed visual descriptions from a medical-purpose knowledge base, for example, “Pneumonia”→ “It is a condition of the lung primarily affecting the small air sacs known as alveolar. It may present with opacities and pleural effusion and it can increase the diagnostic accuracy of lung consolidation”. Note that, such descriptions for the medical terminologies can easily be sourced from either existing educational textbooks or online resources^12^. Despite its simplicity, converting the entities into descriptions is crucial for more reliable and open-vocabulary diagnosis, as it further decomposes the medical entities into visual attributes that are shared by different diseases, encouraging the model to capture a deep understanding of the visual evidence.

To encode the entity, we use the ClinicalBERT [3] as a pre-trained text encoder, to first compute the embedding for the entity description and position, and then adopt a linear MLP to flexibly project the embedding to desired dims:

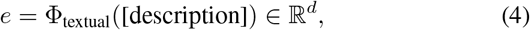

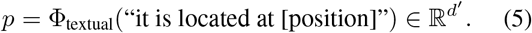

Each triplet has now been converted into {*e, p, l*}, *l* ∈{0, 1} denotes the existence of the entity.

##### Discussion

We have made two major differences compared to the existing visual-language models in computer vision, *First*, the information in medical reports is often more condensed, normally describing the existence of abnormality and their positions in the image, thus, adopting the **filter** operation can avoid unnecessary complexity from grammar, while still retaining most of the useful information in reports. *Second*, entities tend to be medical terminologies that are only understandable to audiences with a medical background, enrich the encoding by visual descriptions can significantly help the model to capture a deep understanding of the visual evidence for diseases, specifically, for seen diseases, such shared visual attributes enable to build the implicit relationship, while for unseen diseases, their visual evidence may have already been well understood by processing the descriptions of the seen ones, as they tend to be shared among diseases.

#### 3.2.3. Fusion Module

After extracting all the entities and their corresponding positions from the reports, we select the top |*Q*| most commonly appearing entities in reports, and compute the textual embeddings for their corresponding descriptions, denoted as an entity set *Q* = {*e*_1_, *e*_2_, …, *e*_|*Q*|_}, and top |*P* | position embeddings as a position set *P* = {*p*_1_, *p*_2_, …, *p*_|*P* |_}. For a certain image, its computed visual representation and the entity set will be passed into a fusion module for alignment, consisting of multiple Transformer Decoder layers. We treat the entity set *Q* as Query, and the image features 𝒱 as Key and Value into the Transformer decoders, the outputs from the fusion module are further fed into two linear MLP layers, one is used for inferring the existence of the entity, and the other generates an embedding to indicate the entity’s spatial position:

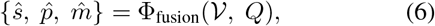

where ŝ ∈ℝ^|*Q*|^ represents the existence prediction for each entity query, and 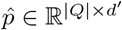 represents the prediction embedding of spatial position for the entities. 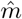 denotes the average of the cross-attention maps sourced from Transformer Decoder layers. The training procedure will be detailed in Sec. 3.3.

##### Discussion

In contrast to the existing approaches [56] that aligns the reports with the entire image, our adopted Transformer decoder enables to compute correspondences between text and image at the patch level. Consequently, the image features 𝒱 are more suitable for downstream segmentation tasks and the average of the cross-attention map in each layers can be used directly for **zero-shot** grounding.

### 3.3. Training

To train the proposed model, we leverage the corresponding triplets, for entities that are not mentioned in the considered report, we simply ignore them while computing loss. For simplicity, the following formulations are based on the assumption that the considered query do have corresponding triplets. In specific, for the existence prediction ŝ, we use binary cross-entropy with the existence label, denoted as ℒ_cls_; to supervise the position prediction for each entity query, we adopt contrastive learning, randomly sample *M* position embeddings from the position set:

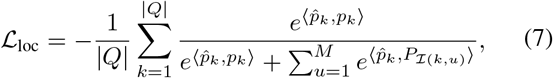

where ⟨·, · ⟩ represents the inner product of two vectors and ℐ (·, ·) is a random index sampling function. *P* is unnormalized in calculation.

The final loss is the sum of the two:

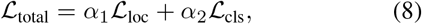

where *α*_1_, *α*_2_ refer to two hyper-parameters controlling the ratio of the two losses, and we set them to be 1.0 by default.

## 4. Experiment

In this section, we start by introducing the dataset used for experiments, *e.g*., pre-training, and various downstream datasets. Then we describe the implementation details and the considered baselines.

### 4.1. Pre-training Dataset

**MIMIC-CXR v2 [19, 28]** consists of over 227k studies of paired image-report data, they are sourced from 65,379 patients at different scanning. Each study can have one or two images (different scan views), totaling 377,110 images.

### 4.2. Datasets for Downstream Tasks

**ChestX-ray14 [50]** contains 112,120 frontal-view X-ray images of 30,805 unique patients, collected from the year of 1992 to 2015 by NIH(National Institutes of Health), with labels of 14 common diseases provided. We split the dataset into 0.8*/*0.1*/*0.1 for train/valid/test.

**RSNA Pneumonia [44]** contains more than 260k frontal-view chest X-rays with corresponding pneumonia opacity masks collected by RSNA (Radiological Society of North America). Commonly, it is treated as a classification tasks [7, 22]. We split the dataset into 0.6*/*0.2*/*0.2 for train/valid/test.

**SIIM-ACR Pneumothorax [1]** contains more than 12k frontal-view chest X-rays with pneumothorax masks collected by SIIM-ACR (Society for Imaging Informatics in Medicine and American College of Radiology). Similarly to RSNA Pneumonia dataset, it can be both used as classification and segmentation tasks. We split the dataset into 0.6*/*0.2*/*0.2 for train/valid/test.

**COVIDx CXR-2 [41] and COVID Rural [12, 47]** aim to evaluate on diagnosing COVID-19. COVIDx CXR-3 contains 29,986 images from 16,648 patients with COVID-19 classification labels. We use it as a classification dataset and split it into 0.7*/*0.2*/*0.1 for train/valid/test. Additionally, we use COVID Rural dataset for COVID-19 segmentation. It contains more than 200 chest X-rays with segmentation masks, and we split it into 0.6*/*0.2*/*0.2 for train/valid/test.

**Edema Severity [8]** contains 6,524 examples from MIMIC-CXR with pulmonary edema severity labels (0 to 3, increasing severity) extracted from the radiology reports. Of these, 141 radiologists were examined by radiologists, and consensus was reached on severity level. It can be seen as a typical fine-grained classification task. We split the dataset into 0.6*/*0.2*/*0.2 for train/valid/test.

### 4.3. Implementation Details

This section details the implementation for architecture, pre-training, zero-shot inference and fine-tuning procedure.

#### Model architecture

As input to the model, images are resized into 224 ×224 ×3. We use the first four layers of ResNet50 [21] as our visual backbone (Φ_visual_), and adopt a MLP layer to transform the output feature dimension into *d* = 256. As a result, the output feature maps from visual encoder is 𝒱∈ ℝ^14×14×256^. On the report side, we extract the entities with a pre-trained text filter, as described in [26], and compute the entity and position embedding with a pre-trained ClinicalBERT [2], its default embedding dim is *d*^′^ = 768. We obtain |*Q*| = 75 entities and |*P* |= 51 positions that most frequently appear in the reports, following [55]. We sample *M* = 7 negative positions for each entity to calculate contrastive loss for training entity position training. In the fusion module, We adopt 4 Transformer Decoder layers with 4 heads in each layer.

#### Pre-training

At this stage, both the filtering operation and language encoding use pre-trained networks, while the visual encoder and fusion module are trained end-to-end on the image-text pairs. We use AdamW [35] optimizer with *lr* = 1 ×10^−4^ and *lr*_warm up_ = 1 ×10^−5^. We train on a GeForce RTX 3090 GPU with batch size 32 for 60 epochs. The first 5 epochs are set for warming up.

#### Inference

At inference time, given a test image, we aim to infer the existence of certain entity / disease, and ground their visual evidence. For those entities that have appeared at training time, we simply adopt the corresponding elements from the entity query set, while for those unseen ones, we replace the entity with a brief description, and treat that as an added query to the model. The existence output can be directly applied for classification and the average cross-attention of different layers in the transformer-based fusion module between specific query and the visual features are used for grounding.

#### Fine-tuning

For the downstream tasks, with large amount of training data, we can fine-tune the model end-to-end, with our pre-trained visual backbone as initialization. Specifically, for image classification task, we adopt ResNet50 [21] and initialize its first four layers with our pre-trained visual encoder. For image segmentation task, we use ResUNet [13] as backbone and initialize its encoder with our pre-trained image encoder.

### 4.4. Baselines

We compare with various existing state-of-the-art medical image-text pre-train methods, namely, ConVIRT [56], GLoRIA [22] and BioViL [7]. Since ConVIRT and GLo-RIA are pre-trained on an in-house dataset, we re-train their models on MIMIC-CXR dataset for fair comparison. For BioViL, we use the officially released models by the authors. For zero-shot setting, we use the prompt as mentioned by BioViL [7]. For fine-tuning, we all use ResNet50 as the visual encoder as described in Sec. 4.3.

### 4.5. Metrics

**AUC** refers to the area under the receiver operating characteristic (ROC) curve, that is commonly used for detection and binary classification tasks.

**F1 and ACC** are used as supplementary metrics for classification tasks. Specifically, F1 comprehensively measures the recall and precision of the model, and ACC is the short of Accuracy. The final binary prediction threshold is chosen to maximise the F1 score. The ACC score is also calculated under this threshold.

**Pointing Game** is used for evaluating the grounding performance. In specific, we extract the region with max response in the output heat-map, for one instance, if the region hit the ground-truth mask, it is considered a positive prediction, otherwise negative. Finally, accuracy can be calculated as the pointing game score.

**Dice and IOU** are commonly used for segmentation tasks. For zero-shot segmentation, we search the segmentation threshold with 0.01 interval for all methods, and report the maximal Dice score for each model.

**Precision and Recall** refer to the detection Precision and Recall. For medical, it is important that lesions are detected even without fine segmentation. Additionally, in some hard cases, especially for the zero-shot setting, Dice and IOU may be too strict to reflect the performance difference. Precision and recall scores can compensate for these. We choose the IOU threshold as 0.1 to calculate the scores.

## 5. Results

In this section, we will report the experimental results. In general, we split the results into two parts: zero-shot setting and fine-tuning setting. In the zero-shot case (Sec. 5.1), we carry out the ablation study and compare it with the other SOTA image-text pre-train methods. We mainly consider classification and segmentation tasks; In the fine-tuning case (Sec. 5.2), we evaluate the model’s transferability by fine-tuning the model with 1%, 10%, and 100% data portion. Additionally, we also add a disease grading downstream task, which can be seen as a fine-grade classification task, showing that our pre-trained model can be transferred to the downstream tasks at ease.

### 5.1. Zero-shot

In this section, we compare our method with the other state-of-the-art methods under zero-shot setting, classification, and grounding. Due to the space limitation, we include the entire ablation study in the supplementary material, referring to it for more details and analysis, and all comparisons here are made using our best model with position contrastive loss and entity description encoder.

#### 5.1.1. Classification

##### Seen Diseases

As shown in Tab. 1, we compare with existing methods on three widely-used datasets, demonstrating consistent performance improvement. Specifically, on pneumonia and pneumothorax datasets, despite the images being collected by different clinics with different diseases, our model improves the AUC score from 0.83 to 0.87 on RSNA pneumonia dataset and from 0.71 to 0.89 on SIIM-ACR pneumothorax dataset, as shown in Tab. 1. This shows that our method can better deal with the multi-center and multi-disease data distribution in medical. While on ChestX-ray14 dataset, we improve the average AUC scores from 0.69 to 0.77, we refer the reader to supplementary material for a more detailed comparison of 14 diseases.

**Table 1.** Comparison with other state-of-the-art methods on zero-shot classification task. AUC, F1 and ACC scores are reported. For ChestX-ray14, the metrics all refer to the macro average on the 14 diseases.

| Dataset<br>Methods | RSNA Pneumonia |  |  | SIIM-ACR Pneumothorax |  |  | ChestX-ray14 |  |  |
| --- | --- | --- | --- | --- | --- | --- | --- | --- | --- |
|  | AUC↑ | F1↑ | ACC↑ | AUC↑ | F1↑ | ACC↑ | AUC↑ | F1↑ | ACC↑ |
| ConVIRT [56] | 0.8042 | 0.5842 | 0.7611 | 0.6431 | 0.4329 | 0.5700 | 0.6101 | 0.1628 | 0.7102 |
| GLoRIA [22] | 0.7145 | 0.4901 | 0.7129 | 0.5342 | 0.3823 | 0.4047 | 0.6610 | 0.1732 | 0.7700 |
| BioViL [7] | 0.8280 | 0.5833 | 0.7669 | 0.7079 | 0.4855 | 0.6909 | 0.6912 | 0.1931 | 0.7916 |
| Ours | **0.8694** | **0.6342** | **0.8002** | **0.8924** | **0.6833** | **0.8428** | **0.7676** | **0.2525** | **0.8619** |

##### Unseen Diseases

In particular, we use covid-19 to evaluate the systems. **Note that**, our considered setting is different from existing approaches, where all entities have been exposed to the model at training time, and prediction can be made by a retrieval-type approach, *i.e*., compute the similarity between the image and the entity embedding by encoding the disease name with a language encoder [7], while we are considering a stricter setting for openset classification. Covid-19 is a new disease that only appeared in 2019, MIMIC-CXR reports collected in the year 2015 do not include any entity of covid-19, thus it requires the system to have the ability to diagnose truly unseen diseases.

As shown in Tab. 2, existing approaches that only rely on disease name struggles to make the correct diagnosis. While with our proposed approach by introducing medical knowledge, *i.e*., using entity descriptions, our methods can understand the complex medical entity descriptions unseen in the training set, and significantly boost the performance 0.66 to 0.74 on AUC and from 0.59 to 0.70 on ACC.

**Table 2.**
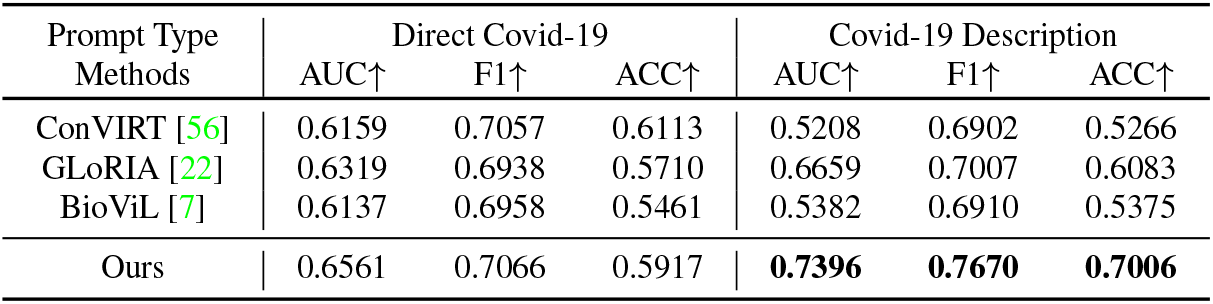
Comparison with other state-of-the-art methods on zero-shot Covid-19 classification task. AUC, F1 and ACC scores are reported. *“Direct covid-19”* refers to directly use “Covid-19” to construct the prompt sentence while *“Covid-19 Description”* refers to replace the name “Covid-19” with its medical description.

| Prompt Type<br>Methods | Direct Covid-19 |  |  | Covid-19 Description |  |  |
| --- | --- | --- | --- | --- | --- | --- |
|  | AUC↑ | F1↑ | ACC↑ | AUC↑ | F1↑ | ACC↑ |
| ConVIRT [56] | 0.6159 | 0.7057 | 0.6113 | 0.5208 | 0.6902 | 0.5266 |
| GLoRIA [22] | 0.6319 | 0.6938 | 0.5710 | 0.6659 | 0.7007 | 0.6083 |
| BioViL [7] | 0.6137 | 0.6958 | 0.5461 | 0.5382 | 0.6910 | 0.5375 |
| Ours | 0.6561 | 0.7066 | 0.5917 | **0.7396** | **0.7670** | **0.7006** |

#### 5.1.2. Grounding

In addition to the plain diagnosis, explainability can be equally critical in healthcare, improving the reliability and trustiness of the machine learning systems. Here, we consider providing explainability by grounding the abnormality in the prediction and compare against the existing approaches. Similarly, we split the diseases into seen and unseen ones, depending on whether their names have appeared in the medical reports. Specifically, “Pneumonia” and “Pneumothorax” are viewed as seen, and “Covid-19” is viewed as unseen. Due to the space limitation, we refer the reader to supplementary material for visualization results.

##### Seen Diseases

We show the results for grounding on RSNA Pneumonia opacity and SIIM-ACR Pneumothorax collapse in Tab. 3. As shown in Tab. 3a, our proposed model surpasses existing approaches on all metrics, for example, we improve the pointing game score from 0.83 to 0.87, the detection Recall from 0.85 to 0.87, the detection precision from 0.50 to 0.64, the IOU from 0.30 to 0.32 and the Dice from 0.44 to 0.46. While on SIIM-ACR dataset (Tab. 3b), the pneumothorax region tends to be thin and narrow, localizing it can often be more challenging than that of opacity grounding [7], we thus only consider pointing game, recall, and precision. Similarly, our method can achieve significantly better performance than prior approaches.

**Table 3.**
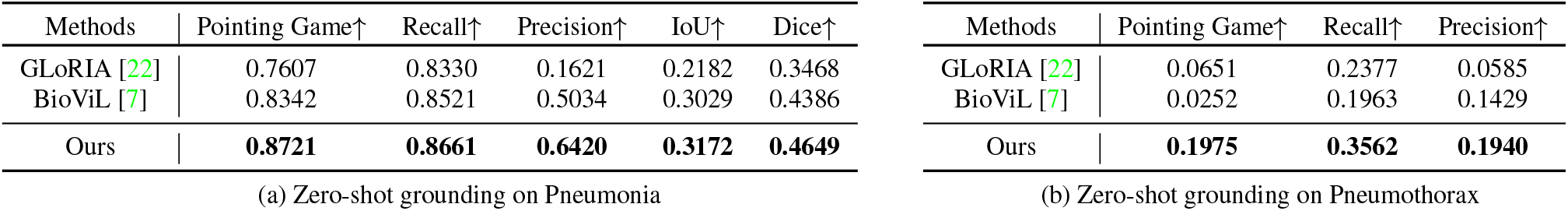
Comparison with other state-of-the-art methods on zero-shot region grounding tasks. (a) shows the results on RSNA Pneumonia dataset. (b) shows the results on SIIM-ACR Pneumothorax dataset. The pneumothorax region tends to be thin and narrow and much more challenging for grounding, we thus only consider pointing game, recall, and precision. Our method can achieve better performance on different metrics, especially on the pointing game score. ConVIRT as the basic method proposed earliest can not realize this function.

##### Unseen Diseases

We also conduct the zero-shot grounding experiment on the unseen disease, namely, Covid-19, as shown in Tab. 4. Our model has shown consistent improvements in all metrics, *e.g*., boosting the pointing game score from 0.40 to 0.58. One observation to be noticed is that, results in Tab. 4 are mostly consistent with those in Tab. 2, *i.e*., better classification results tend to lead to better grounding. Overall, our model with knowledge-enhanced language encoding has facilitated the visual encoder to learn underlying evidence relating to the diseases, therefore, leading to more interpretable representations than prior approaches.

**Table 4.** Comparison with other state-of-the-art methods on zero-shot covid-19 opacity region grounding task. *“Direct covid-19”* refers to directly use “Covid-19” to construct the prompt sentence while *“Covid-19 Description”* refers to replace the name “Covid-19” with its medical description. Our method can achieve better performance on different metrics.

| Prompt Type | Direct covid-19 |  |  |  |  | Covid-19 Description |  |  |  |  |
| --- | --- | --- | --- | --- | --- | --- | --- | --- | --- | --- |
| Methods | Pointing Game↑ | Recall↑ | Precision↑ | IoU↑ | Dice↑ | Pointing Game↑ | AR↑ | AP↑ | IoU↑ | Dice↑ |
| GLoRIA [22] | 0.0364 | 0.2906 | 0.1073 | 0.0645 | 0.1141 | 0.2727 | 0.2821 | 0.1336 | 0.0596 | 0.1075 |
| BioViL [7] | 0.4000 | 0.2564 | 0.2703 | 0.1198 | 0.1967 | 0.1818 | 0.2393 | 0.1637 | 0.0861 | 0.1427 |
| Ours | 0.1818 | 0.1880 | 0.1497 | 0.0747 | 0.1289 | **0.5818** | **0.5214** | **0.4959** | **0.1373** | **0.2278** |

### 5.2. Fine-tuning

In this section, we consider the fine-tuning scenario, with the pre-trained model as initialization, and trained end-to-end on the downstream tasks. We test on three different tasks, namely, classification, segmentation, and grading. In classification and segmentation, the test splits and metrics are the same as in the “zero-shot” section. Grading is a new task we introduce in fine-tuning setting, which can be seen as a fine-grained classification task.

#### 5.2.1. Classification

We experiment on four different datasets, using 1%, 10%, 100% of the data for fine-tuning, that is consistent with the existing work [7, 22, 56]. As shown in Tab. 5, our model has demonstrated substantial improvements in the AUC scores over the existing approaches across all datasets, reflecting that our pre-trained representation is of higher quality than existing models. We refer the readers to the supplementary material for more detailed comparison results.

**Table 5.** Comparison of AUC scores with other state-of-the-art methods on fine-tuning classification task. The macro average of AUC scores on 14 diseases are reported for ChestX-ray14 dataset.

| Dataset<br>Data Portion | Pneumonia |  |  | Pneumothorax |  |  | Covid-19 |  |  | ChestX-ray14 |  |  |
| --- | --- | --- | --- | --- | --- | --- | --- | --- | --- | --- | --- | --- |
|  | 1% | 10% | 100% | 1% | 10% | 100% | 1% | 10% | 100% | 1% | 10% | 100% |
| Scratch | 0.7107 | 0.8150 | 0.8626 | 0.4347 | 0.6120 | 0.6571 | 0.7861 | 0.9162 | 0.9554 | 0.6005 | 0.7365 | 0.7924 |
| ConVIRT [56] | 0.8398 | 0.8562 | 0.8761 | 0.7134 | 0.7826 | 0.9004 | 0.8675 | 0.9541 | 0.9726 | 0.6615 | 0.7658 | 0.8128 |
| GLoRIA [22] | 0.8599 | 0.8666 | 0.8846 | 0.7439 | 0.8538 | 0.9014 | 0.9065 | 0.9381 | 0.9728 | 0.6710 | 0.7642 | 0.8184 |
| BioViL [7] | 0.8233 | 0.8538 | 0.8836 | 0.6948 | 0.7775 | 0.8689 | 0.8989 | 0.9529 | 0.9729 | 0.6952 | 0.7527 | 0.8245 |
| Ours | **0.8731** | **0.8799** | **0.8931** | **0.8527** | **0.9071** | **0.9188** | **0.9224** | **0.9657** | **0.9729** | **0.7721** | **0.7894** | **0.8323** |

#### 5.2.2. Segmentation

In Tab. 6, we conduct fine-tuning experiments on three different diseases for segmentation. We pick 1%, 10%, 100% of the data for fine-tuning. For all three different diseases with different image distributions, our methods surpass the existing state-of-the-art methods by a large margin, especially under the low-data regime.

**Table 6.** Comparison of Dice scores with other state-of-the-art methods on fine-tuning segmentation tasks. Three diseases are reported, and for each disease, three data portions, 1%, 10%, 100% are adopted to show the performance change under different data amounts.

| Diseases<br>Data Portion | Pneumonia |  |  | Pneumothorax |  |  | Covid-19 |  |  |
| --- | --- | --- | --- | --- | --- | --- | --- | --- | --- |
|  | 1% | 10% | 100% | 1% | 10% | 100% | 1% | 10% | 100% |
| Scratch | 0.4347 | 0.6047 | 0.7068 | 0.2133 | 0.3323 | 0.7447 | 0.1481 | 0.2367 | 0.3228 |
| ConVIRT [56] | 0.5706 | 0.6491 | 0.7201 | 0.5406 | 0.6121 | 0.7352 | 0.1995 | 0.2724 | 0.3737 |
| GLoRIA [22] | 0.6555 | 0.6907 | 0.7328 | 0.5673 | 0.5778 | 0.7694 | 0.1889 | 0.2809 | 0.3869 |
| BioViL [7] | 0.6824 | 0.7038 | 0.7249 | 0.6267 | 0.6998 | 0.7849 | 0.2113 | 0.3239 | 0.4162 |
| Ours | **0.7064** | **0.7162** | **0.7579** | **0.6659** | **0.7210** | **0.7937** | **0.2445** | **0.3539** | **0.4399** |

#### 5.2.3. Grading

Besides diagnosis, grading the disease severity level also plays an important role. Here, we adopt our pre-trained features and train them for the multi-class classification task, with 0 to 3 representing different severity levels. As shown in Tab. 7, for each level, the AUC, F1, and ACC scores are calculated as one class against all other ones, for example, 0 vs {1, 2, 3}. Final macro average scores of four levels are computed. On the majority of severity levels, our method can achieve the best results.

**Table 7.** Comparison with other state-of-the-art methods on fine-tuning edema severity grading multi-class classification task. AUC score is reported in the Table. “0,1,2,3” in the table represents the severity level and final macro average scores are reported.

| Methods | 0 |  |  | 1 |  |  | 2 |  |  | 3 |  |  | AVG |  |  |
| --- | --- | --- | --- | --- | --- | --- | --- | --- | --- | --- | --- | --- | --- | --- | --- |
|  | AUC↑ | F1↑ | ACC↑ | AUC↑ | F1↑ | ACC↑ | AUC↑ | F1↑ | ACC↑ | AUC↑ | F1↑ | ACC↑ | AUC↑ | F1↑ | ACC↑ |
| Scratch | 0.7631 | 0.7036 | 0.6738 | 0.5383 | 0.3593 | 0.3223 | 0.6692 | 0.4328 | 0.7012 | 0.8420 | 0.5694 | 0.8770 | 0.7031 | 0.5163 | 0.6436 |
| ConVIRT [56] | 0.8453 | **0.7769** | **0.7793** | 0.6099 | 0.3938 | 0.4629 | 0.7202 | 0.4843 | 0.6445 | 0.9047 | 0.6154 | 0.8809 | 0.7700 | 0.5676 | 0.6919 |
| GLoRIA [22] | 0.8304 | 0.7577 | 0.7520 | 0.6208 | 0.3991 | 0.4922 | 0.7339 | 0.4958 | **0.7037** | **0.9246** | **0.6667** | 0.9102 | 0.7774 | 0.5798 | 0.7145 |
| BioViL [7] | 0.8034 | 0.7378 | 0.7148 | 0.6035 | 0.3912 | 0.4570 | 0.6860 | 0.4497 | 0.6777 | 0.9229 | 0.6500 | 0.9160 | 0.7540 | 0.5572 | 0.6914 |
| Ours | **0.8502** | 0.7646 | 0.7539 | **0.6641** | **0.4140** | **0.5392** | **0.7605** | **0.5266** | 0.7031 | 0.8845 | 0.6250 | **0.9160** | **0.7898** | **0.5826** | **0.7280** |

## 6. Conclusion

In this paper, we introduce a novel medical knowledge enhanced VLP model. First, we propose a report filter to extract useful medical entities with more useful supervision signals, simplifying complex raw reports with minimal information loss. Second, we translate entities into detailed medical descriptions and embed them with a text encoder enabling the network to understand complex medical expert-level knowledge. Finally, a transformer-based structure is proposed to do local region alignment. In experiments, We evaluate our method on different datasets under various settings. Our method shows strong zero-shot classification and grounding abilities, even facing unseen diseases. Besides, in fine-tuning setting, our method still outperforms state-of-the-art methods significantly, showing the superiority of our method.

## Data Availability

All data produced in the present study are available upon reasonable request to the authors

## Supplementary

### A. The Entity Description Base and Position Set

Tab. 8 shows the descriptions we used to translate different entities. We have kept 75 entities in query set *Q*, following [55]. “Tail abnorm obs” entity represents some tailed entities and “exluded obs” represents some entities useless for diagnosis. The last “covid-19” description row is only referred to for inference since it does not appear in pre-train reports.

**Table 8.**
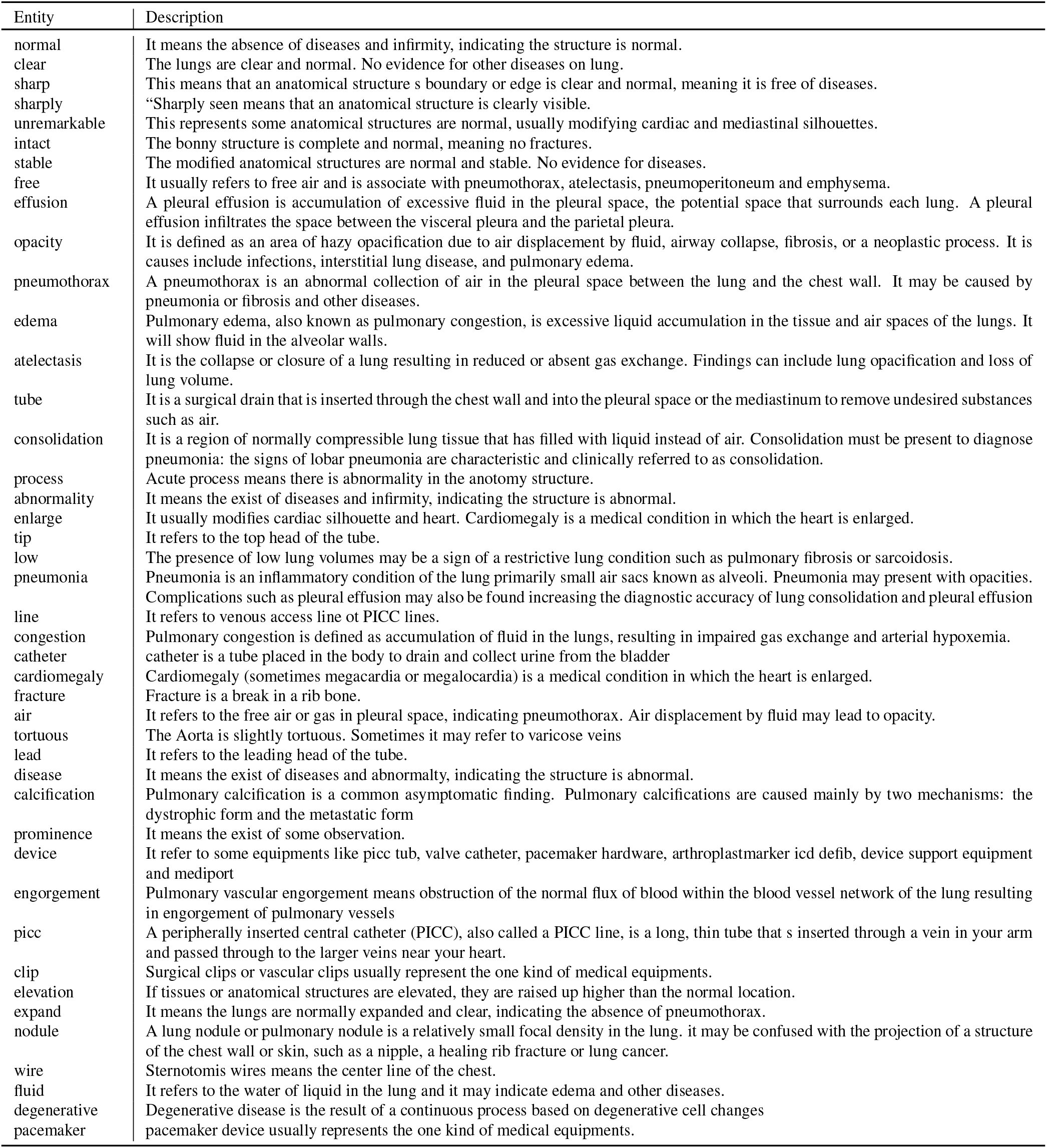
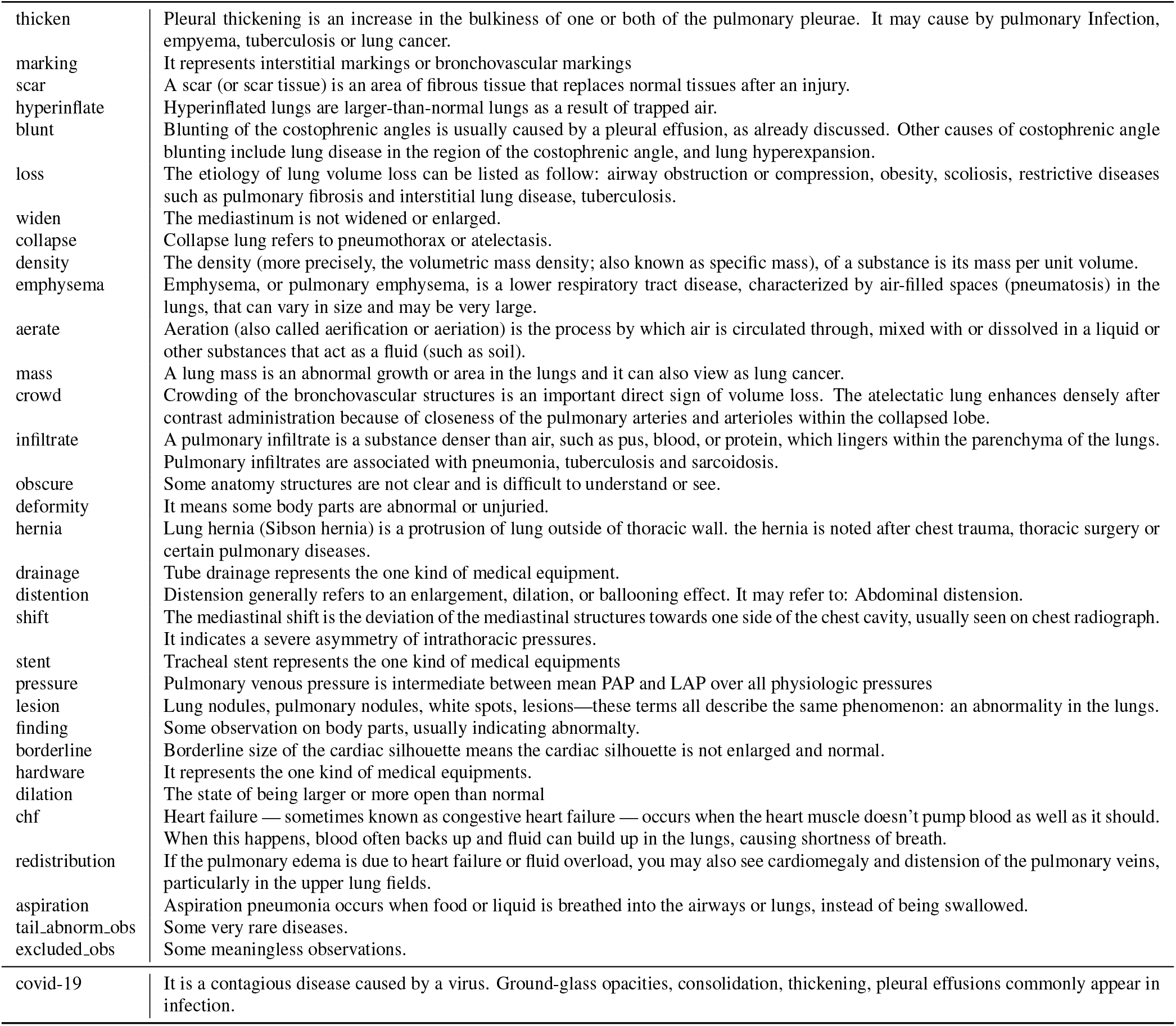
The Entity description used for translate single entity name. The description can be easily found from the open website.

Additionally, we keep 51 positive positions, following [55], to form the position set *P*, as {*trachea, left_hilar, right_hilar, hilar_unspec, left_pleural, right_pleural, pleural_unspec, heart_size, heart_border, left_diaphragm, right_diaphragm, diaphragm_unspec, retrocardiac, lower_left_lobe, upper_left_lobe, lower_right_lobe_middle_right_lobe, upper_right_lobe, left_lower_lung, left_mid_lung, left_upper_lung_left_apical_lung, left_lung_unspec, right_lower_lung, right_mid_lung, right_upper_lung_right_apical_lung, right_lung_unspec, lung_apices, lung_bases, left_costophrenic_right_costophrenic, costophrenic_unspec, cardiophrenic_sulcus, mediastinal, spine_clavicle, rib, stomach, right_atrium, right_ventricle, aorta, svc, interstitium, parenchymal, cavoatrial junction, cardiopulmonary, pulmonary, lung_volumes, unspecified, other*}. “Other” is used to reprepresnt some tailed positions.

### B. Details of Fusion Module

In the transformer-based fusion module, the queries are first passed through a self-attention layer and then followed by a multi-head attention layer between the modified queries and image features. In each head, the image features are processed by a linear key head and a linear value head as key embeddings and value embeddings independently. The value is weighted-added based on the attention map, which is calculated by the soft-max dot product of the keys and queries. Finally, a feed-forward network gathers the vector of different heads resulting in the output of this layer. The output vector of the former layer is considered as the entity query vector of the next layer. In formulation, if denoting 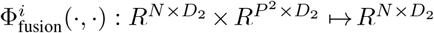 as the i-th layer, the procedure is expressed as:

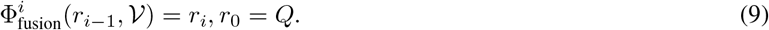

### C. Ablation Study

Our final method mainly contains three key parts, transformer-based fusion module, position location contrastive loss (PosCL), and entity description encoder (DE). We gradually remove the modules to analyze their effectiveness. “w/o (DE)” refers to removing DE module and “w/o (PosCL+DE)” refers to only maintaining the fusion module with basic CE loss. Tab. 9 and Tab. 10 shows the quantitative results.

**Table 9.**
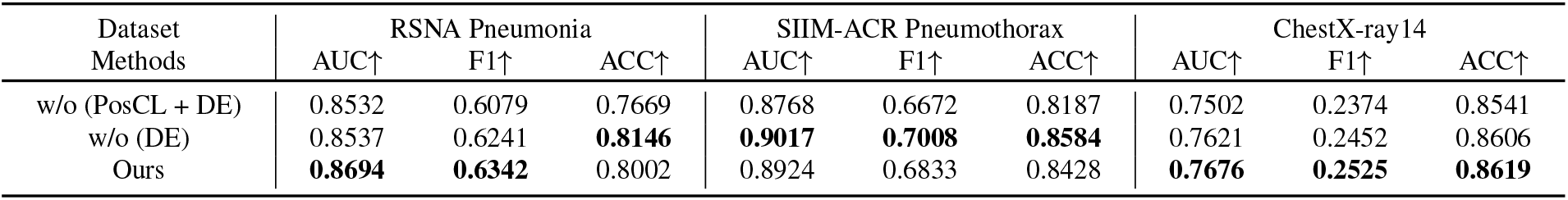
Ablation study on zero-shot classification task. AUC, F1 and ACC scores are reported. For ChestX-ray 14, the metrics all refer to the macro average on the 14 diseases.

**Table 10.** Ablation study on zero-shot grounding tasks. (a) shows the results on RSNA Pneumonia dataset. (b) shows the results on SIIM-ACR Pneumothorax dataset.

| Methods | Pointing Game↑ | Recall↑ | Precision↑ | IoU↑ | Dice↑ | Methods | Pointing Game↑ | Recall↑ | Precision↑ |
| --- | --- | --- | --- | --- | --- | --- | --- | --- | --- |
| w/o (PosCL + DE) | 0.7979 | **0.8961** | 0.4036 | 0.2783 | 0.4230 | w/o (PosCL + DE) | 0.1786 | 0.3151 | 0.1336 |
| w/o (DE) | 0.8424 | 0.8226 | **0.6520** | 0.3118 | 0.4610 | w/o (DE) | **0.2080** | 0.3178 | 0.1711 |
| Ours | **0.8721** | 0.8661 | 0.6420 | **0.3172** | **0.4649** | Ours | 0.1975 | **0.3562** | **0.1940** |

(a) Zero-shot grounding on Pneumonia
(b) Zero-shot grounding on Pneumothorax

#### Transformer Decoder

The lines about “w/o (PosCL + DE)” in tables demonstrate the performance of the basic model modified only by base CE loss. This model can exceed many former methods. This indicates the complex syntax will hurt the network to capture the useful entities significantly and our filtering operation combined with medical NER can greatly relieve the problem.

#### Position Contrastive Loss

The PosCL can significantly help the network to ground the abnormalities. As shown in the results by adding PosCL the classification results can be further improved, e.g., from 0.75 to 0.76 on AUC in ChestX-ray14 dataset. Besides classification, location contrastive loss can bring more gain in grounding. These results show position is another vital element in reports especially for grounding tasks. Our filtered triplets can conclude and clean the reports with little information loss and make the network learn the report information more straightforward.

#### Entity Description Encoder

By adding entity descriptions, we want to realize two goals. *First*, in addition to just learning from the image-report data, the network can actively learn the relationship between different entities based on the entity descriptions. As shown in tables, adding descriptions in most scenarios can help the network better understand the entity and bring gain to the final metric scores. *Second*,the description encoder enables our model to **handle openset new diseases**. Since the entity list is a close set during pre-training, our method will be only able to handle the seen diseases without DE, while, with a description encoder, our method can handle unseen diseases and understand complex medical disease knowledge.

### D. Detailed results on ChestX-ray14

We further show the detailed performance of 14 different diseases on ChestX-ray14 dataset. Tab. 11 shows the results on the zero-shot setting. Our method can exceed the former methods for most diseases. The radar Fig. 3Y shows more visually how our model compares with other solutions under the zero-shot setting. Our method can exceed the former methods for most diseases. Under 100% fine-tuning settings, we achieved similarly excellent results as shown in Tab. 12.

**Table 11.** Comparison with other state-of-the-art methods on zero-shot ChestX-ray 14 diseases classification task. For each disease, AUC score is reported and the macro average AUC score is also reported. We use the first three letters to represent one disease but for “pneumonia” and “pneumothorax” we use the first two and the last letters.

| Methods | Ate. | Car. | Eff. | Inf. | Mas. | Nod. | Pna. | Pnx. | Con. | Ede. | Emp. | Fib. | Thi. | Her. | AVG |
| --- | --- | --- | --- | --- | --- | --- | --- | --- | --- | --- | --- | --- | --- | --- | --- |
| ConVIRT [56] | 0.4533 | 0.4601 | 0.7262 | 0.6238 | 0.6790 | 0.6322 | 0.6097 | 0.6698 | 0.6855 | 0.7699 | 0.4701 | 0.5293 | 0.6098 | 0.6220 | 0.6101 |
| GLoRIA [22] | 0.6680 | 0.7647 | 0.7975 | 0.6159 | 0.6722 | 0.5293 | 0.6755 | 0.4785 | 0.7306 | 0.8212 | 0.6033 | 0.5104 | **0.6721** | 0.7144 | 0.6610 |
| BioViL [7] | 0.5026 | 0.6328 | 0.7914 | 0.5791 | 0.7029 | 0.6126 | 0.6866 | 0.7516 | 0.7455 | 0.8533 | 0.7136 | 0.6751 | 0.6560 | 0.7692 | 0.6909 |
| w/o (PosCL + DE) | 0.7131 | 0.8100 | 0.8635 | **0.6361** | 0.7776 | 0.6740 | 0.6903 | 0.8124 | 0.7915 | 0.8869 | 0.7480 | **0.6780** | 0.6429 | 0.7784 | 0.7502 |
| w/o (DE) | 0.7420 | 0.8270 | **0.8663** | 0.6336 | 0.7867 | **0.6974** | **0.7238** | 0.8310 | 0.8037 | 0.8887 | 0.7865 | 0.6715 | 0.5414 | 0.8691 | 0.7621 |
| ours | **0.7506** | **0.8299** | 0.8636 | 0.6280 | **0.7885** | 0.6947 | 0.7236 | **0.8361** | **0.8079** | **0.8888** | **0.7950** | 0.6511 | 0.5783 | **0.9097** | **0.7676** |

**Table 12.** Comparison with other state-of-the-art methods on fine-tuning ChestX-ray 14 diseases classification task. For each disease, AUC score is reported and the macro average AUC score is also reported. We use the first three letters to represent one disease but for “pneumonia” and “pneumothorax” we use the first two and the last letters.

| Methods | Ate. | Car. | Eff. | Inf. | Mas. | Nod. | Pna. | Pnx. | Con. | Ede. | Emp. | Fib. | Thi. | Her. | AVG |
| --- | --- | --- | --- | --- | --- | --- | --- | --- | --- | --- | --- | --- | --- | --- | --- |
| Scratch | 0.7835 | 0.8116 | 0.8563 | 0.6537 | 0.7788 | 0.6912 | 0.7004 | 0.8561 | 0.8090 | 0.8869 | 0.8564 | 0.7534 | 0.7454 | 0.9106 | 0.7924 |
| ConVIRT [56] | 0.8012 | 0.8360 | 0.8511 | 0.6613 | 0.8004 | 0.7490 | 0.6998 | 0.8666 | 0.8079 | 0.9023 | 0.9014 | 0.7933 | 0.7468 | 0.9627 | 0.8128 |
| GLoRIA [22] | 0.8263 | 0.8326 | 0.8596 | **0.6641** | 0.8179 | 0.7348 | 0.7104 | 0.8452 | 0.8129 | 0.8977 | **0.9310** | 0.7886 | 0.7608 | 0.9750 | 0.8184 |
| BioViL [7] | 0.8185 | 0.8543 | 0.8607 | 0.6660 | 0.8302 | 0.7633 | 0.7090 | 0.8595 | **0.8287** | 0.9031 | 0.9251 | 0.7912 | 0.7638 | 0.9696 | 0.8245 |
| ours | **0.8291** | **0.8594** | **0.8719** | 0.6565 | **0.8382** | **0.7647** | **0.7378** | **0.8807** | 0.8275 | **0.9083** | 0.9224 | **0.7977** | **0.7784** | **0.9796** | **0.8323** |

**Figure 3.**
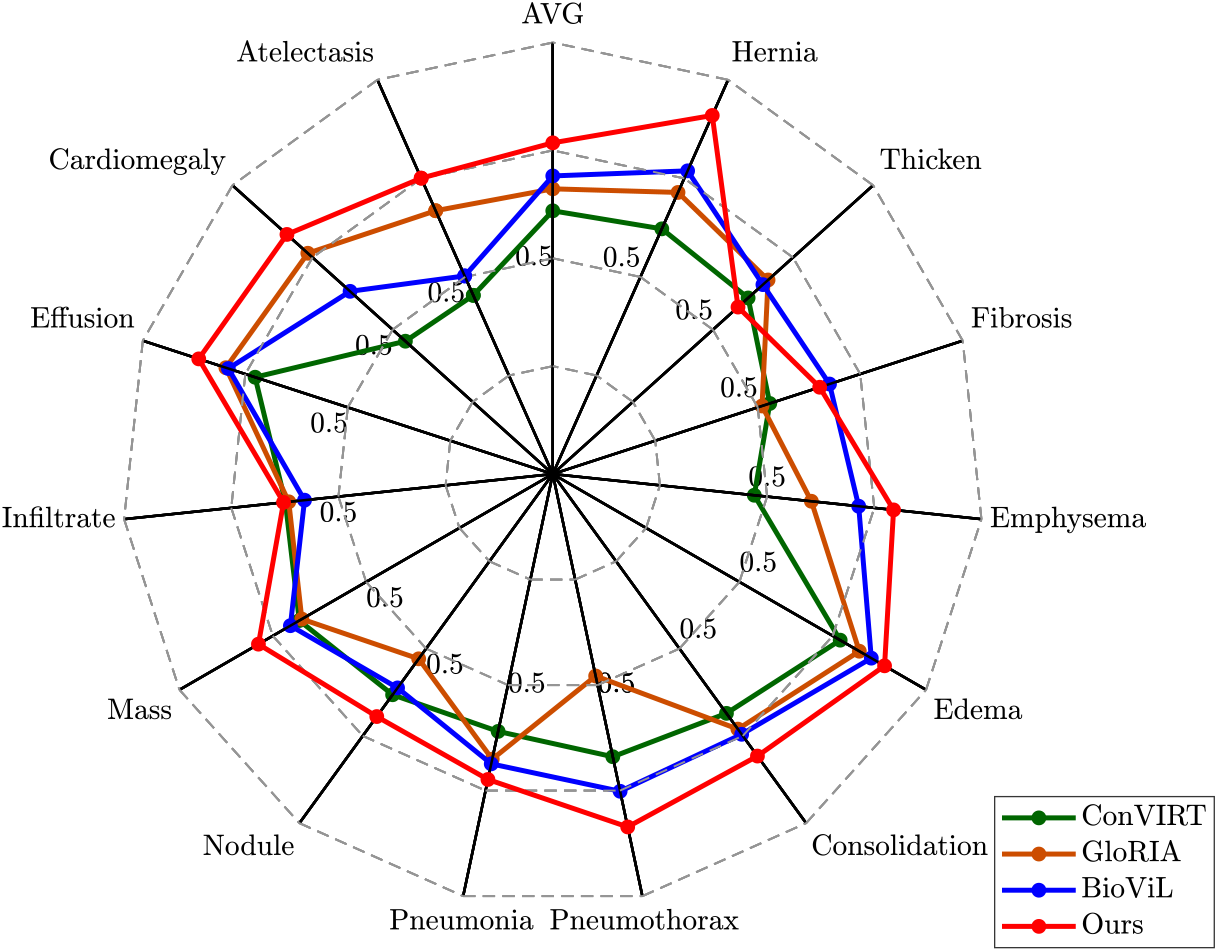
The radar figure of our method and other methods of ChestX-ray14 14 diseases. AUC scores are reported and, as shown, our method exceeds the previous state-of-the-art on most diseases.

### E. Visualization Results

Fig. 4 shows visualization results of our model on zero-shot grounding task. As shown in figure, the ground truth of “Pneumonia” is given by bounding box and generally related to a large area region. Thus the metrics on this are higher than other two datasets. Our network captures its regions very well. For “Pneumothorax”, its abnormality pattern is different from other diseases, which aim to capturing the collapsed part of the lung, rendering darker areas on the images rather than brighter opacity. Its ground-truth masks are generally thin and narrow while our network can still highlight its location. For “Covid-19”, its image textual was similar to “Pneumonia”, but since this is a totally new disease, grounding its regions is much more challenging. It requires the model to build relationships between them based on their complex definition and symptoms. The visualization results suggest that our model successfully achieve this, supporting that, for other unseen diseases, our model can also understand their complex descriptions.

**Figure 4.**
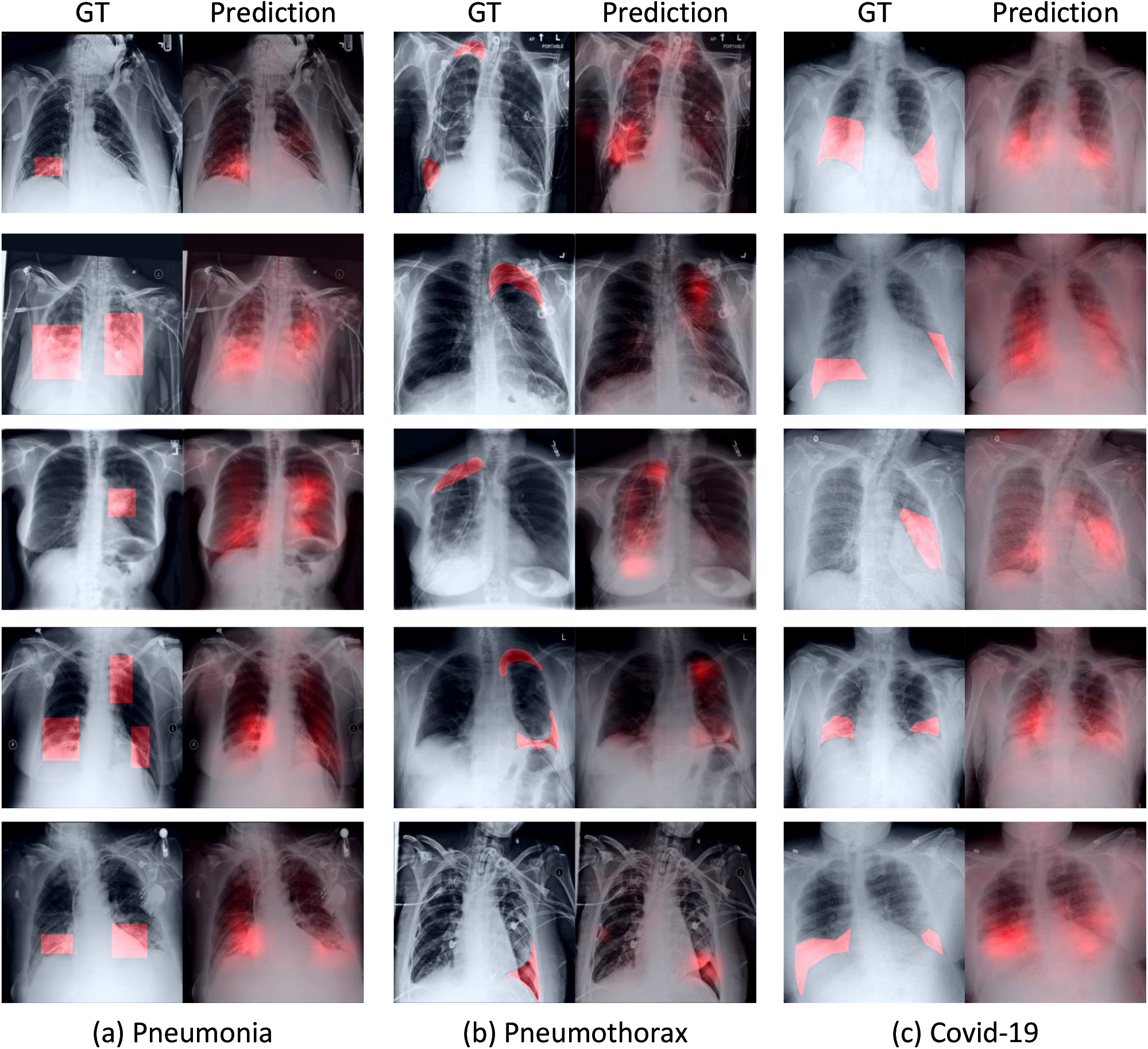
The visualization of zero-shot grounding results of our method. Each column represents the results on one disease and the left in it is the ground-truth and right is the heatmap predication of our model. The brighter the red on the figure, the more likely the model considering this region to be associated with abnormalities.

## Footnotes

1 Wikipedia https://en.wikipedia.org/wiki/Wiki

2 UMLS [6] https://www.nlm.nih.gov/research/umls/index.html

